# Characteristics of SARS-CoV-2 Testing for Rapid Diagnosis of COVID-19 during the Initial Stages of a Global Pandemic

**DOI:** 10.1101/2020.12.23.20231589

**Authors:** Jennifer L. Guthrie, Allison J. Chen, Dalton R. Budhram, Kirby Cronin, Adriana Peci, Paul Nelson, Gustavo V. Mallo, George Broukhanski, Michelle Murti, Anna Majury, Tony Mazzulli, Vanessa G. Allen, Samir N. Patel, Julianne V. Kus, Vanessa Tran, Jonathan B. Gubbay

## Abstract

Accurate SARS-CoV-2 diagnosis is essential to guide prevention and control of COVID-19. From January 11 – April 22, 2020, Public Health Ontario conducted SARS-CoV-2 testing of 86,942 specimens collected from 80,354 individuals, primarily using real-time reverse-transcription polymerase chain reaction (rRT-PCR) methods. We analyzed test results across specimen types and for individuals with multiple same-day and multi-day collected specimens. Nasopharyngeal compared to throat swabs had a higher positivity (8.8% vs. 4.8%) and an adjusted estimate 2.9 C_t_ lower (SE=0.5, *p*<0.001). Same-day specimens showed high concordance (98.8%), and the median C_t_ of multi-day specimens increased over time. Symptomatic cases had rRT-PCR results with an adjusted estimate 3.0 C_t_ (SE=0.5, *p*<0.001) lower than asymptomatic/pre-symptomatic cases. Overall test sensitivity was 84.6%, with a negative predictive value of 95.5%. Molecular testing is the mainstay of SARS-CoV-2 diagnosis and testing protocols will continue to be dynamic and iteratively modified as more is learned about this emerging pathogen.

## INTRODUCTION

As severe acute respiratory syndrome coronavirus 2 (SARS-CoV-2) spreads globally, specific, rapid, and reliable diagnostic tests are essential to minimise spread through timely contact investigation and isolation of infected individuals. Rapid detection of SARS-CoV-2 by real-time reverse-transcription polymerase chain reaction (rRT-PCR) is currently the standard testing method for coronavirus disease 2019 (COVID-19) diagnosis(1). rRT-PCR has been used to detect SARS-CoV-2 from upper respiratory specimens such as nasopharyngeal swabs (NPS), throat swabs (TS), and saliva(2,3), lower respiratory specimens such as sputum, bronchoalveolar lavage, and in some cases non-respiratory specimens(4). Early studies detected higher viral loads in respiratory over non-respiratory specimens(5), with NPS currently the recommended specimen type(6). Viral loads are typically highest early in the course of illness and decrease after approximately 10-14 days following symptom onset, as reflected in rRT-PCR results at or near the limit of detection(6–8).

Diagnostic challenges have arisen as SARS-CoV-2 has been detected across a range of disease manifestations, including pre-symptomatic, asymptomatic, and atypical presentations(5,9,10). Presently, there is limited information regarding the association between patient demographics, clinical presentation, viral load and SARS-CoV-2 detection, particularly with respect to serial testing over time. More studies are needed to inform the optimal protocol for SARS-CoV-2 diagnostic testing.

Here, we examine SARS-CoV-2 molecular-based test performance characteristics and summarize case-level data related to COVID-19 diagnosis. We analyze rRT-PCR test results across different specimens collected over single and multiple days for individuals. As testing protocols have been dynamic and iteratively modified among laboratories, our study findings will inform the further development and refinement of molecular-based testing and related testing algorithms.

## MATERIALS and METHODS

### Study Setting and Design

As the provincial reference laboratory, Public Health Ontario (PHO) conducts a large proportion of SARS-CoV-2 testing for Ontario, Canada. During the initial stage of the pandemic, all specimens collected from individuals across the province were submitted to PHO. After March 11, 2020, testing in Ontario was decentralized to increase capacity, and a number of hospital and private laboratories implemented SARS-CoV-2 diagnostic testing. Included in this study were all specimens received for SARS-CoV-2 routine testing by PHO from January 11, 2020 through April 22, 2020.

### Specimen Collection

Specimens submitted to PHO included upper respiratory specimens such as nasopharyngeal, nasal, and throat swabs, lower respiratory specimens (e.g. sputum, bronchoalveolar lavage, lung tissue), and non-respiratory specimens (e.g. blood, stool). Upon collection, swabs were placed in transport media and sent to PHO for processing(11). Initially, collection of 2 specimens from different upper respiratory sites was recommended, which was later revised to a single specimen due to high demand for swabs.

### Data Collection

Demographic, epidemiological and clinical data were abstracted from PHO test requisitions. To reflect changing knowledge regarding COVID-19, these requisitions were updated on several occasions (see **Supplementary Figure S1** for key changes in testing guidelines). Information was recorded at the time of specimen collection and included demographics, healthcare setting, self-reported symptoms(12), and reasons for testing such as travel history and contact with a known or probable COVID-19 case. Specimen type was also recorded. It should be noted that initial requisition versions provided a nasopharyngeal checkbox but not a separate nasal swab option, therefore NPS described herein may represent a compilation of nasal, mid-turbinate and NPS specimens.

### Molecular-based Detection of SARS-CoV-2

Several methods of SARS-CoV-2 detection were implemented over the first few months of the pandemic as newly developed tests and platforms became available. Initially, a nested end-point PCR was used targeting the SARS-CoV-2 RNA-dependent RNA polymerase (RdRp) gene(13)(**Supplementary Appendix I**) followed by Sanger sequencing using standard methods. Early specimens were also sent to Canada’s National Microbiology Laboratory (NML) for secondary confirmation and supplementary testing using various NML-developed molecular-based assays. Next, an rRT-PCR laboratory developed test (LDT) was used for detection of the *Sarbecovirus* subgenus envelope (E) gene and/or SARS-CoV-2 RdRp gene based on Corman et al.(14) primer and probe sequences. Interpretation of the LDT rRT-PCR cycle threshold (C_t_) was as follows: C_t_ ≤38 was categorized as detected (positive), 38.1 – 39.9 was indeterminate, and a C_t_ ≥40 was not detected (negative). Initially, the testing algorithm required detection of both targets for laboratory confirmation. This was later revised to single target detection for COVID-19 diagnosis using the E-gene target alone. As of mid-March, to increase laboratory capacity, a commercial rRT-PCR assay, cobas® SARS-CoV-2 (Roche Diagnostics, Germany) was introduced, which includes E-gene and ORF1ab targets. Specimens tested using the cobas® SARS-CoV-2 rRT-PCR assay were reported as detected (positive) or not detected (negative); the manufacturer does not include an indeterminate category. Uninterpretable results for any test method were reported as invalid. For further details of RNA extraction, PCR protocols and test result definitions see **Supplementary Appendix I**. C_t_ values reported herein were based on E-gene test results, as this was the common target used in both the LDT and cobas® SARS-CoV-2 rRT-PCR assays.

### Data Analysis

Demographic and clinical information were summarised at an individual-level. For persons with multiple same-day collected specimens, any positive result was considered a positive for that day. Collection day was counted from the first date (day 1) of specimen(s) collected for an individual. Missing collection dates were substituted with laboratory login dates as the mean difference between these was 1 day (data not shown). A testing episode was defined as the overall result for an individual on a collection day.

### Statistical Analysis

All statistical analyses were performed in R (v4.0.2). Categorical variables were summarized using frequency and percentage with comparison of positive and negative test results for study variables measured using the chi-square (χ^2^) test. Non-normally distributed continuous variables were summarized using median and interquartile range (IQR) with differences assessed using the non-parametric Mann-Whitney U test, and Wilcoxon signed-rank test for paired samples. A generalised linear regression model was used to examine C_t_ values with standard error (SE) of NPS and TS adjusted for symptom status, age and gender. Unless otherwise stated indeterminate results were counted as positives.

## RESULTS

### Study Sample Characteristics

A total of 86,942 specimens with valid test results were collected from 80,354 individuals and received at PHO from January 11, 2020 through April 22, 2020 (**Figure 1**). This represents 47% of the more than 184,000 SARS-CoV-2 laboratory tests completed in Ontario during this period(15). Overall, 6,834 individuals (8.5%) had a positive SARS-CoV-2 test result. The median age of individuals with a positive test was 72 years (IQR 50–87), significantly higher (*p*<0.001) than individuals with a negative test, median age 54 years (IQR 36–77)(**Table 1**). The largest proportion of cases was observed in the 80– 89 year age group (22.5%) and amongst females (57.1%). Reasons for testing were recorded at the time of specimen collection, and of all positive cases, 4,650 (68.0%) had COVID-19 symptoms, 747 (10.9%) had international travel history, and 646 (9.5%) had contact with a known or probable COVID-19 case. Of these reasons, case contact had a higher likelihood for association with a positive result. Characteristics of persons with invalid results (*n*=145) can be seen in **Supplementary Table S1**.

**Table 1.**
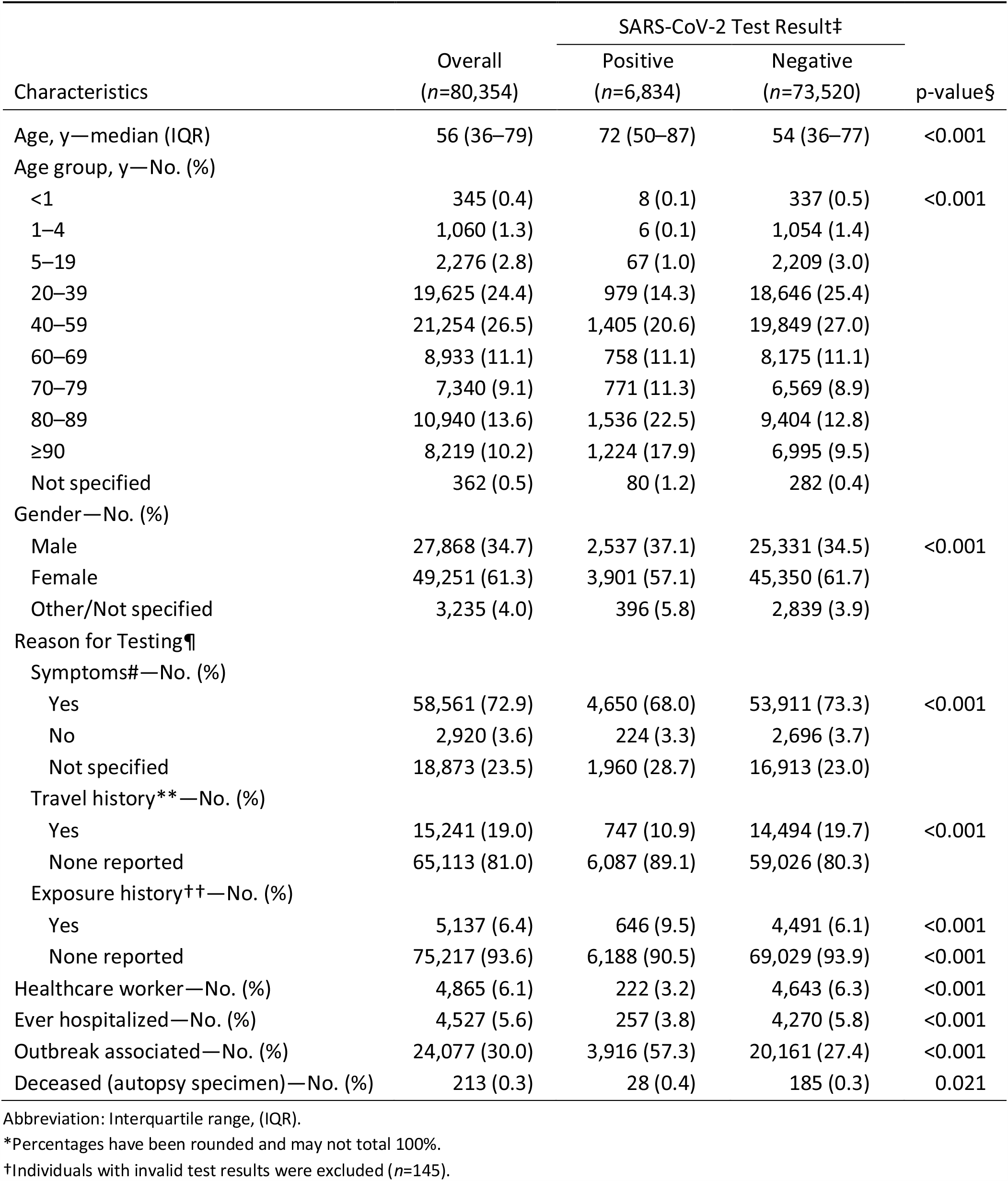
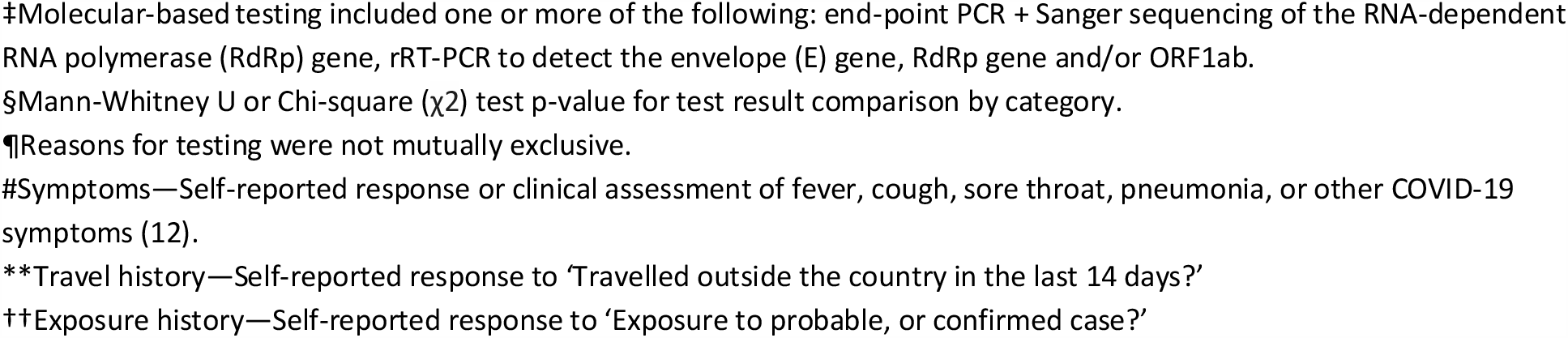
Characteristics of 80,354 individuals with specimens received at Public Health Ontario January 11, 2020 – April 22, 2020 for SARS-CoV-2 diagnostic testing.*†

| Characteristics | Overall<br>(n=80,354) | SARS-CoV-2 Test Result‡ |  | p-value§ |
| --- | --- | --- | --- | --- |
|  |  | Positive<br>(n=6,834) | Negative<br>(n=73,520) |  |
| Age, y—median (IQR) | 56 (36–79) | 72 (50–87) | 54 (36–77) | <0.001 |
| Age group, y—No. (%) |  |  |  |  |
| <1 | 345 (0.4) | 8 (0.1) | 337 (0.5) | <0.001 |
| 1–4 | 1,060 (1.3) | 6 (0.1) | 1,054 (1.4) |  |
| 5–19 | 2,276 (2.8) | 67 (1.0) | 2,209 (3.0) |  |
| 20–39 | 19,625 (24.4) | 979 (14.3) | 18,646 (25.4) |  |
| 40–59 | 21,254 (26.5) | 1,405 (20.6) | 19,849 (27.0) |  |
| 60–69 | 8,933 (11.1) | 758 (11.1) | 8,175 (11.1) |  |
| 70–79 | 7,340 (9.1) | 771 (11.3) | 6,569 (8.9) |  |
| 80–89 | 10,940 (13.6) | 1,536 (22.5) | 9,404 (12.8) |  |
| ≥90 | 8,219 (10.2) | 1,224 (17.9) | 6,995 (9.5) |  |
| Not specified | 362 (0.5) | 80 (1.2) | 282 (0.4) |  |
| Gender—No. (%) |  |  |  |  |
| Male | 27,868 (34.7) | 2,537 (37.1) | 25,331 (34.5) | <0.001 |
| Female | 49,251 (61.3) | 3,901 (57.1) | 45,350 (61.7) |  |
| Other/Not specified | 3,235 (4.0) | 396 (5.8) | 2,839 (3.9) |  |
| Reason for Testing¶ |  |  |  |  |
| Symptoms#—No. (%) |  |  |  |  |
| Yes | 58,561 (72.9) | 4,650 (68.0) | 53,911 (73.3) | <0.001 |
| No | 2,920 (3.6) | 224 (3.3) | 2,696 (3.7) |  |
| Not specified | 18,873 (23.5) | 1,960 (28.7) | 16,913 (23.0) |  |
| Travel history**—No. (%) |  |  |  |  |
| Yes | 15,241 (19.0) | 747 (10.9) | 14,494 (19.7) | <0.001 |
| None reported | 65,113 (81.0) | 6,087 (89.1) | 59,026 (80.3) |  |
| Exposure history††—No. (%) |  |  |  |  |
| Yes | 5,137 (6.4) | 646 (9.5) | 4,491 (6.1) | <0.001 |
| None reported | 75,217 (93.6) | 6,188 (90.5) | 69,029 (93.9) | <0.001 |
| Healthcare worker—No. (%) | 4,865 (6.1) | 222 (3.2) | 4,643 (6.3) | <0.001 |
| Ever hospitalized—No. (%) | 4,527 (5.6) | 257 (3.8) | 4,270 (5.8) | <0.001 |
| Outbreak associated—No. (%) | 24,077 (30.0) | 3,916 (57.3) | 20,161 (27.4) | <0.001 |
| Deceased (autopsy specimen)—No. (%) | 213 (0.3) | 28 (0.4) | 185 (0.3) | 0.021 |

Abbreviation: Interquartile range, (IQR).
\*Percentages have been rounded and may not total 100%.
†Individuals with invalid test results were excluded (n=145).
‡Molecular-based testing included one or more of the following: end-point PCR + Sanger sequencing of the RNA-dependent RNA polymerase (RdRp) gene, rRT-PCR to detect the envelope (E) gene, RdRp gene and/or ORF1ab.
§Mann-Whitney U or Chi-square ( $\chi^2$ ) test p-value for test result comparison by category.
¶Reasons for testing were not mutually exclusive.

**Figure 1.**
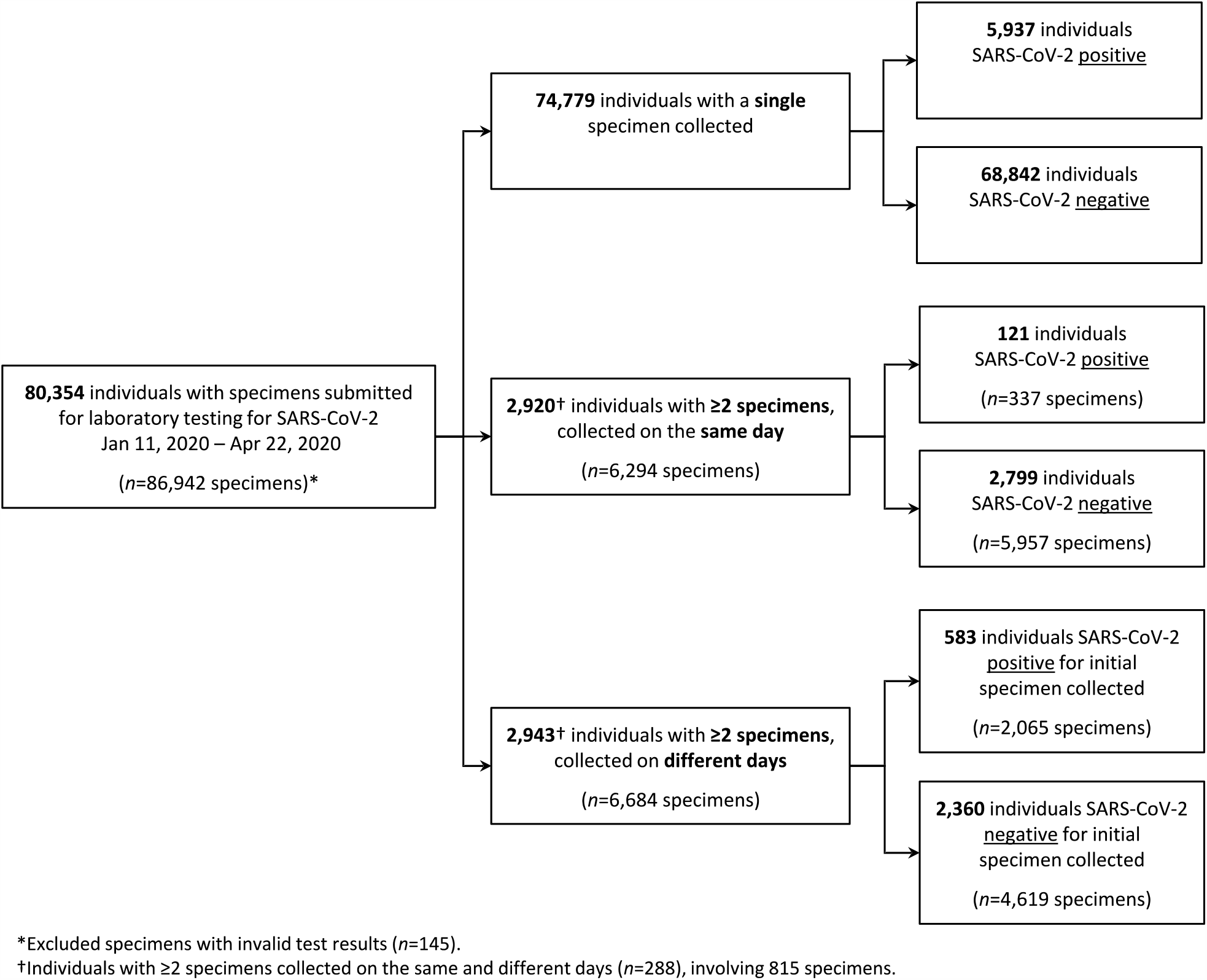
SARS-CoV-2 molecular-based testing results for individuals with specimens submitted to Public Health Ontario, January 11, 2020 – April 22, 2020. Individuals were considered positive if any specimen had rRT-PCR detection of the envelope (E) gene, RNA-dependent RNA polymerase (RdRp) gene, ORF1ab and/or end-point PCR + Sanger sequencing of the RdRp gene. Indeterminate results were counted as positive for the purpose of this analysis (*n*=19).

The number of specimens received for testing increased as the pandemic evolved, with more than 4,000 specimens tested per day on several days in April (**Figure 2**). Supply chain issues limiting the availability of reagents in late-March are reflected in the decreased number of specimens collected and tested at that time. As testing continued, we observed an increase in the proportion of outbreak-associated specimens, and by the study end date, more than 50% of positive specimens were linked to an outbreak investigation (**Figure 2**).

**Figure 2.**
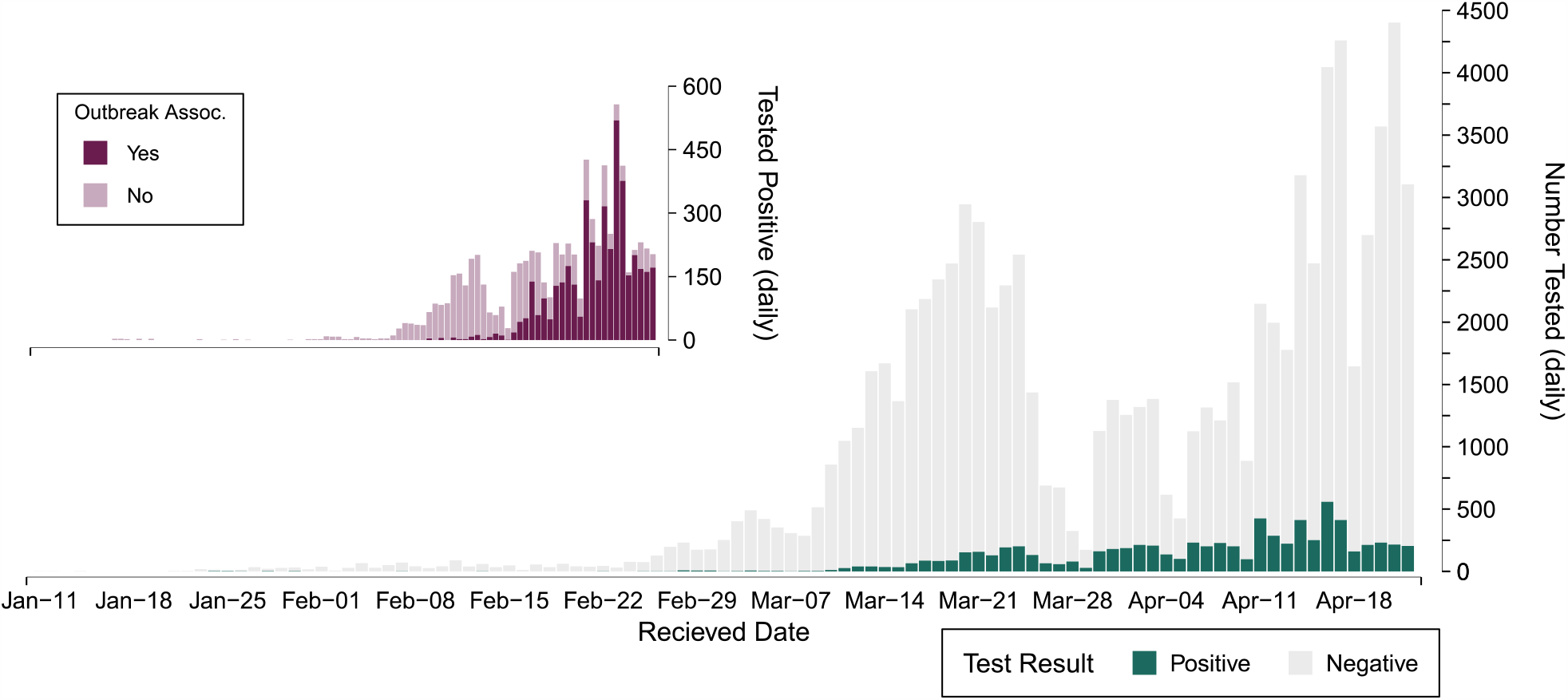
Daily SARS-CoV-2 testing results for specimens received at Public Health Ontario (January 11 – April 22, 2020), with inset plot of daily positives coloured by the number of outbreak-associated specimens over the same time period.

### Test Performance Characteristics

The majority (99.0%, *n*=86,071) of specimens were tested with a single assay, the largest number of which were tested using the LDT rRT-PCR assay (*n*=56,928) or cobas® SARS-CoV-2 assay (*n*=29,141) (**Supplementary Table S3**). Concordant results were observed for 831 (95.4%) specimens tested using multiple assays. Of the 23 specimens with discordant results, 19 had negative end-point PCR results, and where rRT-PCR had also been performed a median of C_t_ 36.2 (IQR 34.4–36.9) was observed. For the remaining 4 discordants, 3 represented negative NML-referred test results and 1 negative LDT result. Details of the discordant results and other test related data can be found in **Supplementary Appendix I**.

### Comparison of Specimen Types

Of 86,942 specimens included in the study, 7,538 (8.7%) tested positive for SARS-CoV-2. The median C_t_ across all assays for the first positive specimen of each case was 24.3 (IQR 19.6–30.4). Upper respiratory specimens accounted for nearly all specimens submitted (98.8%, *n*=85,889), the majority (94.0%) of which were NPS (*n*=81,765)(**Table 2**). For respiratory specimens, NPS had the highest proportion of positives (8.8%) compared to TS (4.8% positive). Furthermore, initial NPS-positive specimens were estimated to have a C_t_ 2.9 (SE=0.5, *p*<0.001) lower than initial TS-positive specimens after adjusting for symptom status, age and gender (**Table 3**). Lower respiratory specimens such as bronchoalveolar lavage, lung tissue, and sputum, accounted for 0.5% of all specimens. Of the 422 lower respiratory specimens, 38 (9.0%) tested positive with a median C_t_ value of 27.0 (IQR 22.7–33.4). Non-respiratory specimens such as cerebral spinal fluid, autopsy tissues, and whole blood, were uncommon (0.01% of all specimens, *n*=13), and only a single non-respiratory specimen (stool – the only one submitted during the study period) tested positive (C_t_ 31.9).

**Table 2.** SARS-CoV-2 molecular-based* test results by specimen type. Specimens received January 11, 2020 – April 22, 2020 (*n*=86,942) and tested at Public Health Ontario with valid results.

| Specimen Source | Specimens Tested<br>*n* (%)† | SARS-CoV-2 Test Result*† |  |
| --- | --- | --- | --- |
|  |  | Positive<br>*n* (%) | Negative<br>*n* (%) |
| Upper Respiratory |  |  |  |
| Nasopharyngeal‡ | 81,765 (94.0) | 7,222 (8.8) | 74,543 (91.2) |
| Throat | 4,064 (4.7) | 197 (4.8) | 3,867 (95.2) |
| Other | 60 (0.1) | 1 (1.7) | 59 (98.3) |
| Lower Respiratory |  |  |  |
| Sputum | 269 (0.3) | 33 (12.3) | 236 (87.7) |
| Bronchoalveolar lavage | 94 (0.1) | 1 (1.1) | 93 (98.9) |
| Other | 59 (0.1) | 4 (6.8) | 55 (93.2) |
| Non-Respiratory | 13 (<0.1) | 1 (7.7) | 12 (92.3) |
| Not specified | 618 (0.7) | 79 (12.8) | 539 (87.2) |

\*Molecular-based testing included one or more of the following: end-point PCR + Sanger sequencing of the RNA-dependent RNA polymerase (RdRp) gene, rRT-PCR to detect the envelope (E) gene, RdRp gene and/or ORF1ab. Percentages have been rounded and may not total 100%.

**Table 3.** Multiple linear regression analysis of factors influencing cycle threshold for the envelope (E) gene. Analysis includes the initial positive specimen for each individual and uses a complete-case analysis strategy (*n*=4,540).

|  | *n* (%) | Unadjusted Estimate<br>(SE) | Adjusted* Estimate<br>(SE) | p-value |
| --- | --- | --- | --- | --- |
| Specimen type |  |  |  |  |
| Nasopharyngeal swab | 4,391 (96.7) | Reference | Reference |  |
| Throat swab | 143 (3.2) | 2.7 (0.5) | 2.9 (0.5) | <0.001 |
| Lower respiratory | 6 (0.1) | 2.6 (2.6) | 3.0 (2.6) | 0.257 |
| Symptoms |  |  |  |  |
| Yes | 4,325 (95.3) | Reference | Reference |  |
| No | 215 (4.7) | 2.9 (0.5) | 3.0 (0.5) | <0.001 |
| Age, y | 4,540 (100.0) | 0.003 (0.004) | 0.002 (0.004) | 0.591 |
| Gender |  |  |  |  |
| Male | 1,783 (39.3) | Reference | Reference |  |
| Female | 2,531 (55.7) | 0.4 (0.2) | 0.5 (0.2) | 0.018 |
| Other/Not specified | 226 (5.0) | 0.02 (0.5) | 0.1 (0.5) | 0.834 |

Abbreviation: SE, standard error.
\*Adjusted for all variables – specimen type, symptoms, age, gender.

### Detection of SARS-CoV-2 with Repeated Sampling

We analyzed the results of 5,575 individuals with multiple specimens submitted to provide insight to same-day and multi-day sampling (**Figure 1**). The majority of individuals (86.8%, *n*=4,837) had 2 specimens collected (**Supplementary Table S7**). Amongst individuals with multiple specimens, 2,632 (45.4%) had all specimens collected on a single day, and 2,874 (49.6%) had one or more specimen collected over a 1–60 day interval. There were 288 (5.0%) individuals with both multiple same-day and multi-day specimens collected.

#### Same-day Collected Specimens

We identified 2,920 individuals with multiple specimens collected on the same day (**Figure 1**), resulting in 3,024 multi-specimen testing episodes. Concordant results between same-day specimens were observed for 2,987 (98.8%) multi-specimen testing episodes, of which 105 (3.5%) were positive for all specimens and 2,882 (96.5%) were negative for all specimens. Discordant results between same-day specimens were seen for 37 of 3,024 (1.2%) testing episodes, of these 35 were discordant positive/negative results, 1 positive/indeterminate and 1 negative/indeterminate result. Specimen types and test results are categorized in **Supplementary Table S8**.

Within the 105 concordant same-day positive results, there were 26 NPS pairs. For these pairs the overall median C_t_ was 24.8 (IQR 19.0–32.0), and median C_t_ difference between NPS paired swabs was 1.8 (IQR 0.8–3.7)(**Figure 3**). Additionally, there were 72 episodes of same-day NPS/TS pairs with concordant results, for which the TS generally had a higher C_t_ (median difference 3.0 C_t_, IQR -0.6–6.3, *p*<0.001)(**Figure 3**).

**Figure 3.**
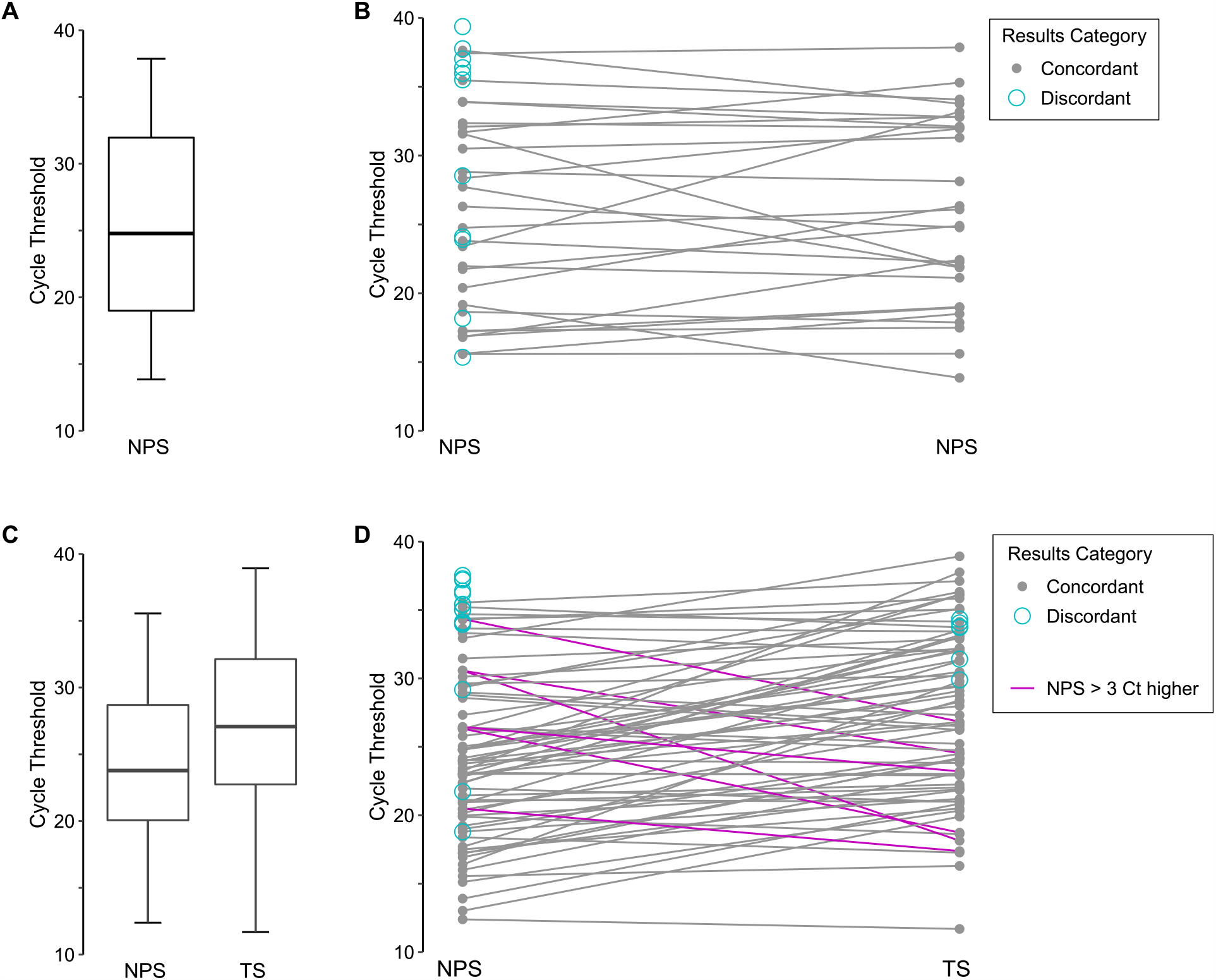
Comparison of envelope (E) gene rRT-PCR cycle threshold (C_t_) values for individuals with multiple same-day specimens collected. C_t_ values are analyzed by episodes for which there were multiple nasopharyngeal swabs (NPS) collected, or a combination of throat swab (TS) and NPS. C_t_ is plotted by results category which indicates that the same-day specimens were both positive (concordant), or had a positive specimen and negative specimen (discordant). **(A)** Boxplot with median C_t_ for all same-day collected nasopharyngeal swabs (NPS) with concordant results, **(B)** C_t_ across same-day collected NPS pairs, **(C)** boxplot with median C_t_ for same-day collected NPS and throat specimens with concordant results, **(D)** C_t_ across same-day collected NPS and throat specimen pairs. Paired NPS + throat specimens for which the NPS was >3 C_t_ higher than the throat specimen are shown as pink lines.

Examination of discordant results revealed 12 NPS specimen pairs (median C_t_ 35.7, IQR 24.1– 37.2), 16 NPS-positive/TS-negative pairs (median C_t_ 35.0, IQR 34.0–36.3), 7 NPS-negative/TS-positive pairs (median C_t_ 33.8, IQR 32.0–34.0), 1 NPS-positive/TS-indeterminate pair (NPS C_t_ 36.8, TS C_t_ unavailable) and 1 NPS-positive/sputum-negative pair (C_t_ 31.2). One individual had 5 specimens collected on the same day with discordant results – 2 TS-negative specimens while the NPS, an unspecified respiratory swab, and an unspecified specimen type were positive. These specimens were taken 6 days after the initial collection of positive specimens (NPS and TS) for this individual. Ultimately, if a single NPS specimen had been collected for the 105 concordant positive pairs and the 37 discordant pairs, up to 13.4% (19/142) of positive (including indeterminate) testing episodes may have been reported negative. However, 4 of these were individuals with previous positive results and did not represent missed diagnoses. Collection of a single TS would have resulted in 16.2% (16 of 99 with a TS collected) negative testing episodes, 11 of which had previous positive results.

#### Multi-day Collected Specimens

A total of 2,943 (3.7%) individuals had specimens collected on more than one day. The time course and result of each of these tests can be found in **Supplementary Figure S3**. Examining the demographics of individuals with multi-day specimens we noted that these persons were older (median 73 years, IQR 52–87) compared to individuals with specimens collected at a single time point (median 55 years, IQR: 36–78; *p*<0.001), and a larger proportion were hospitalized at initial specimen collection (13.1%, 387/2,943) compared to those with single time-point collection (5.1%, 3,910/77,411; *p<*0.001). Furthermore, a larger proportion (39.2%, 1,153/2,943) of individuals with multi-day specimen collection were linked to an outbreak investigation at initial collection compared to those with single day specimens (29.1%, 22,495/77,411; *p*<0.001).

Of the individuals with multi-day collected specimens, 199 initially tested positive and later tested negative over a period of 1 to 37 days (median 15 days, IQR 15–18). The time course of individual tests can be viewed in **Figure 4A**. We also examined C_t_ over time for persons with multi-day collection and observed a steady increase in median C_t_ (**Figure 4B**). For asymptomatic individuals a higher median C_t_ can be seen for the initial specimen collected compared to symptomatic persons. While this observation was based on a small number of asymptomatic individuals (*n*=6) with multi-day testing, this finding was further supported by multiple linear regression using all individuals testing positive in the study (persons with single and multi-day specimens). Asymptomatic persons had an estimated 3.0 C_t_ (SE=0.5, *p*<0.001) higher compared to symptomatic persons, adjusting for specimen type, age and gender (**Table 3**).

**Figure 4.**
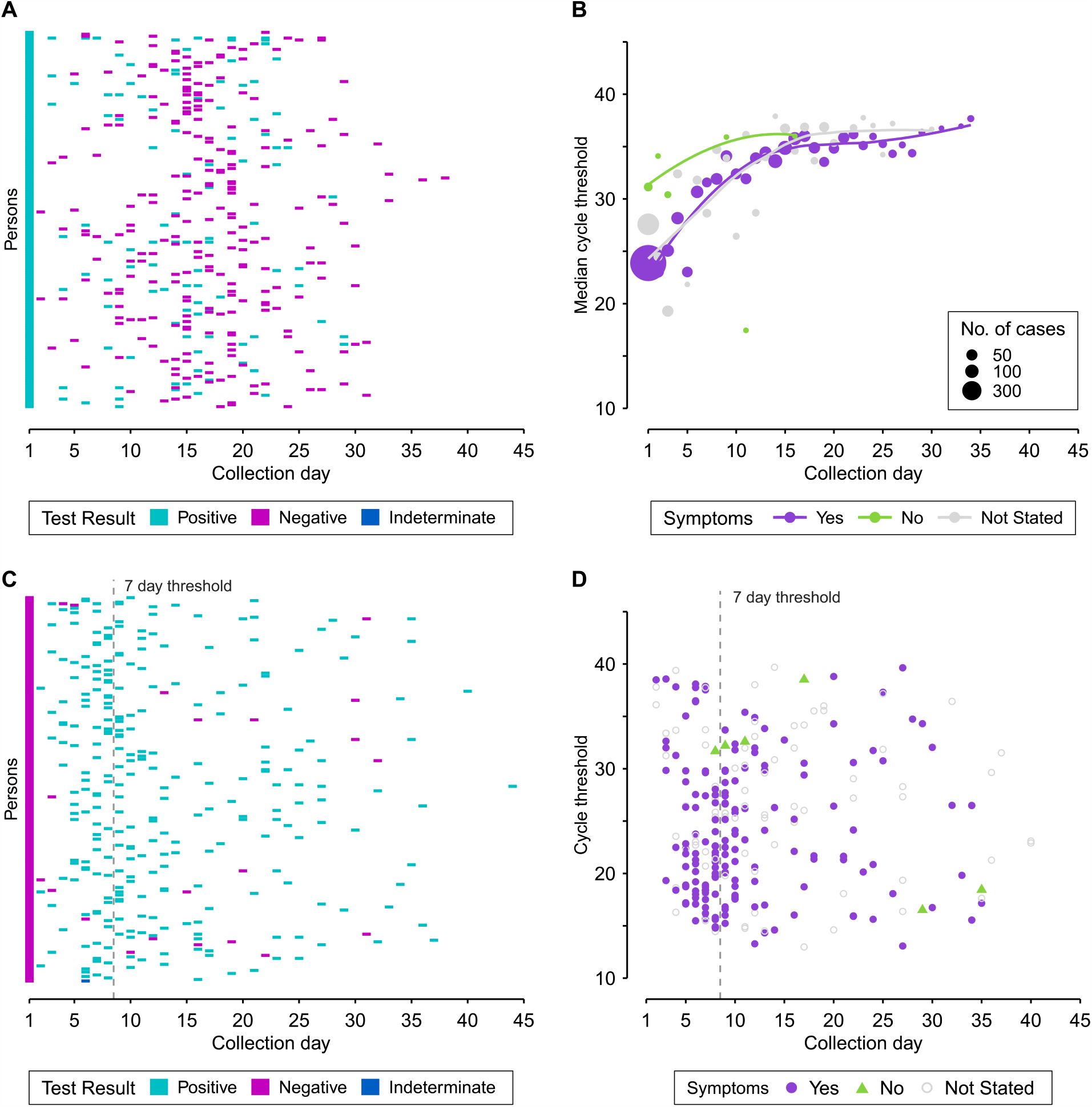
Molecular-based test results for individuals with specimens submitted to Public Health Ontario for SARS-CoV-2 testing, January 11 – April 22, 2020. Any positive result for an individual with multiple specimens collected on the same-day is displayed as positive, and indeterminate where there was an indeterminate result and no positive. **(A)** Pattern of results for individuals initially testing positive (day 1) with a subsequent negative result at a later date (*n*=199). Each individual is represented horizontally across collection days. **(B)** Median cycle threshold (C_t_) by specimen collection day for those with day 1 positive results (*n*=578). Trend lines were fitted using locally estimated scatterplot smoothing (LOESS), with the asymptomatic curve excluding the day 11 outlier. **(C)** Pattern of results for individuals initially testing negative (day 1) with a subsequent positive result at a later date (*n*=255). A threshold 7 days following initial negative result differentiates episodes. **(D)** C_t_ values by day of specimen collection for the first positive result obtained for an individual with a negative result on initial testing. Testing included one or more of the following: end-point PCR + Sanger sequencing of RNA-dependent RNA polymerase (RdRp) gene, rRT-PCR to detect the envelope (E) gene, RdRp gene and/or ORF1ab.

Of the 2,360 persons initially testing negative, 255 (10.8%) individuals had a subsequent positive result (**Figure 4C**). The median number of days to a positive result was 8 (IQR 6–15) with a median C_t_ of 24.0 (IQR 18.9–30.6) for the subsequent positive. Under the assumption that a negative result followed by a positive within 7 days represented an initial false negative finding, we calculated a test sensitivity of 84.6% and negative predictive value of 95.5% (**Table 4**). Beyond 7 days was considered a discrete disease episode as viral load typically peaks within this period and the existing literature suggests viral load and infectivity is diminished within a week following symptom onset(3,16).

**Table 4.** rRT-PCR testing results* for individuals with specimens collected on more than one day (*n*=2,943) and received by Public Health Ontario, January 11, 2020 – April 22, 2020.

| Initial Test Result | COVID-19 Status† |  | Total |
| --- | --- | --- | --- |
|  | Positive | Negative |  |
| Positive | 583 | – | 583 |
| Negative | 106 | 2,254 | 2,360 |

\*Molecular-based testing included one or more of the following: end-point PCR + Sanger sequencing of the RNA-dependent RNA polymerase (RdRp) gene, rRT-PCR to detect the envelope (E) gene, RdRp gene and/or ORF1ab.
†COVID-19 status was based on a review of all test results within the 7 day period following the initial test result. Persons with any positive result within the 7 day period were considered positive, and negative where all results were negative in this time period.

We investigated further the characteristics of this negative-to-positive test result group. The majority (97.2%, *n*=212/218) of specimens collected were NPS, and of the 174 individuals with symptom status documented at initial testing, 162 (93.1%) were symptomatic. On the subsequent positive test, 102 (63.0%) remained symptomatic, with 39 of these individuals testing positive within 7 days, and 63 testing positive more than 7 days following initial negative test (**Supplementary Table S9**). Of note, 10 of 12 asymptomatic individuals on initial negative testing reported symptoms at subsequent positive testing, averaging 7.2 days (SD±2.1) to positive result. Interestingly, 184 (72.2%) individuals with negative-to-positive results were tested in relation to an outbreak. On their follow-up positive test, 40 additional individuals had specimens submitted in association with an outbreak where none was identified on their initial negative test. In the group of negative-to-positive individuals the majority (85.9%, 219/255) did not have contact to a known/probable case indicated on initial or subsequent testing. However, 18 persons without previous exposure indicated contact to a known/probable case on subsequent positive test, with a median of 12.1 days (SD±11.8) from negative result. Additionally, we reviewed C_t_ values for positive results following an initial negative, and no clear pattern was observed in relation to symptom status or time since collection of the initial negative specimen (**Figure 4D**).

## DISCUSSION

Molecular-based testing using nucleic acid amplification techniques are currently the gold standard for SARS-CoV-2 detection. These methods can provide high sensitivity and quick turnaround time. Rapid spread of the virus globally created an unprecedented need for increased SARS-CoV-2 diagnostic testing, and consequently a need to establish how and when to best use these tests, and better interpret the results. In this study, we summarized patient data related to laboratory-based COVID-19 diagnosis for 6,834 cases and described diagnostic testing characteristics for more than 86,000 individuals and specimens tested by PHO during the first few months of the pandemic.

We analyzed the characteristics of individuals tested for COVID-19, and as per previous reports(17,18), persons testing positive were more likely to be older, with more than 60% of cases aged ≥60 years. Early in the pandemic testing priority was given for persons of any age having travelled to specific regions(19). However, recommendations were adjusted as the pandemic evolved with COVID-19 symptoms representing the principal reason for testing overall(20). Of persons reporting symptoms, only 1 in 8 tested positive, which highlights the non-specific nature of COVID-19 symptoms and a major challenge of this pandemic(21).

Our findings of higher positivity and lower C_t_ values for NPS relative to TS is consistent with the higher clinical sensitivity of NPS reported in the literature(22,23). In accordance with current recommendations for, and widespread availability of, NPS, most specimens in this study were upper respiratory. While PHO tested lower respiratory and non-respiratory specimens, these specimen types represented a small proportion of all specimens tested. Interestingly, sputum had a comparatively high positivity (12.9%), similar to previous reports(24), and may represent an underutilized specimen type in situations where it is warranted. However, interpretations of results for rare specimen types should be made with caution as the reasons for the choice of specimen type collected were not available, and these individuals may have had a higher pre-test probability(25).

Where multiple specimens were collected on the same day, nearly all results were concordant.

In the small number of cases with discordant results, C_t_ values were often high and the discordance likely represents a level of virus beyond the rRT-PCR limit of detection. This may be related to the time of sampling in the disease course (C_t_ is lowest within the first week following symptom onset(6)), or poor sample collection(25,26). Where specimen type differed for same-day collection, NPS were more often positive, similar to other studies(2,27) further supporting the use of NPS as the preferred specimen type.

In this study, specimens were collected on different days for nearly 3,000 individuals, with C_t_ values increasing over disease course and a higher median C_t_ for asymptomatic persons compared to those with symptoms, similar to previous findings(6,7,28). It should be noted that there have also been reports of similar viral load between symptomatic and asymptomatic individuals(6,29). Asymptomatic individuals without epidemiological risk factors (i.e. low pre-test probability) may in some cases represent false positive results. Distinguishing between a true asymptomatic case and a high C_t_ false positive is a significant diagnostic and public health challenge. Particularly as questions remain regarding the extent of asymptomatic/pre-symptomatic transmission in the community. False negatives also have the potential to complicate efforts in controlling SARS-CoV-2, as these individuals – believed not to be infected – are not directed to isolate and may unknowingly facilitate virus dissemination. In our study, we identified 255 persons with an initial negative test that at a later date tested positive. Of these, nearly three-quarters were initially tested in relation to an outbreak. These individuals may have been truly negative initially – tested as part of screening efforts during an outbreak, and were later exposed due to continued association with the outbreak setting.

Our study has several limitations. First, reported sensitivity and NPV should be interpreted with caution as our analysis does not compare two distinct tests, and instead uses an individual’s collective test results within a 7 day period (“true positive” result) as a comparison for their initial test result. Therefore, sensitivity was dependent on the time period used as a discrete episode. Current literature reports a 5 day mean incubation with viral shedding a few days before symptom onset. Thus, we defined an episode as 7 days following initial specimen collection, and found our calculation of test sensitivity to be consistent with the literature(2); however, our NPV estimate was higher than other reports(26). As it is more likely for persons with high pre-test probability to be retested, there may be a bias towards an underestimation of the true sensitivity. The nature of this method does not allow for calculation of specificity and PPV. Second, a limitation of many COVID-19 studies at this time, including this one, is that recommendations regarding who should be tested changed over the study period and may have impacted which individuals were prioritized for testing. Thus, our results may not be representative of all cases, particularly asymptomatic or hospitalized individuals, and may not be generalizable to all locations. Last, drawing conclusions regarding the asymptomatic/pre-symptomatic infected population is difficult within this study population owing to limited data on test requisitions. For example, timing of sample collection in disease course (symptom onset date was often not captured, and is not possible for persons without symptoms) and reason(s) for testing, limited information regarding exposure, and lack of follow-up testing in most cases.

In conclusion, the present study contributes to the rapidly expanding body of knowledge regarding SARS-CoV-2 clinical diagnostics. Our results support existing estimates of test sensitivity while providing further insights into the relationship between C_t_ and patient-related factors, as well as the variability between specimen types, multiple same-day and repeat testing. Diagnostics has a key role to play in the pandemic, contributing to policy and practice regarding individual- and population-level public health measures used to limit spread of the virus.

## Data Availability

Patient data cannot be shared publicly for ethical and legal reasons, as public availability would compromise patient privacy. Researchers who meet criteria for access to confidential data can contact the PHO Privacy Officer.

## ACKNOWLEDGMENTS

We gratefully acknowledge the staff of Virus Detection and Molecular Diagnostics, Public Health Ontario Laboratory, for diagnostic testing of SARS-CoV-2 specimens. We also acknowledge with thanks to the National Microbiology Laboratory (NML), Winnipeg, MB for providing confirmatory and supplementary testing early in the pandemic.

## ETHICS

The Public Health Ontario Ethics Review Board has determined that this project did not require research ethics committee approval, as it describes analyses that were completed at Public Health Ontario Laboratory as part of routine clinical respiratory testing during the first wave of the COVID-19 pandemic in Ontario and are therefore considered public health practice.

