## Supplemental Materials for "Characteristics of SARS-CoV-2 Testing for Rapid Diagnosis of COVID-19 during the Initial Stages of a Global Pandemic"

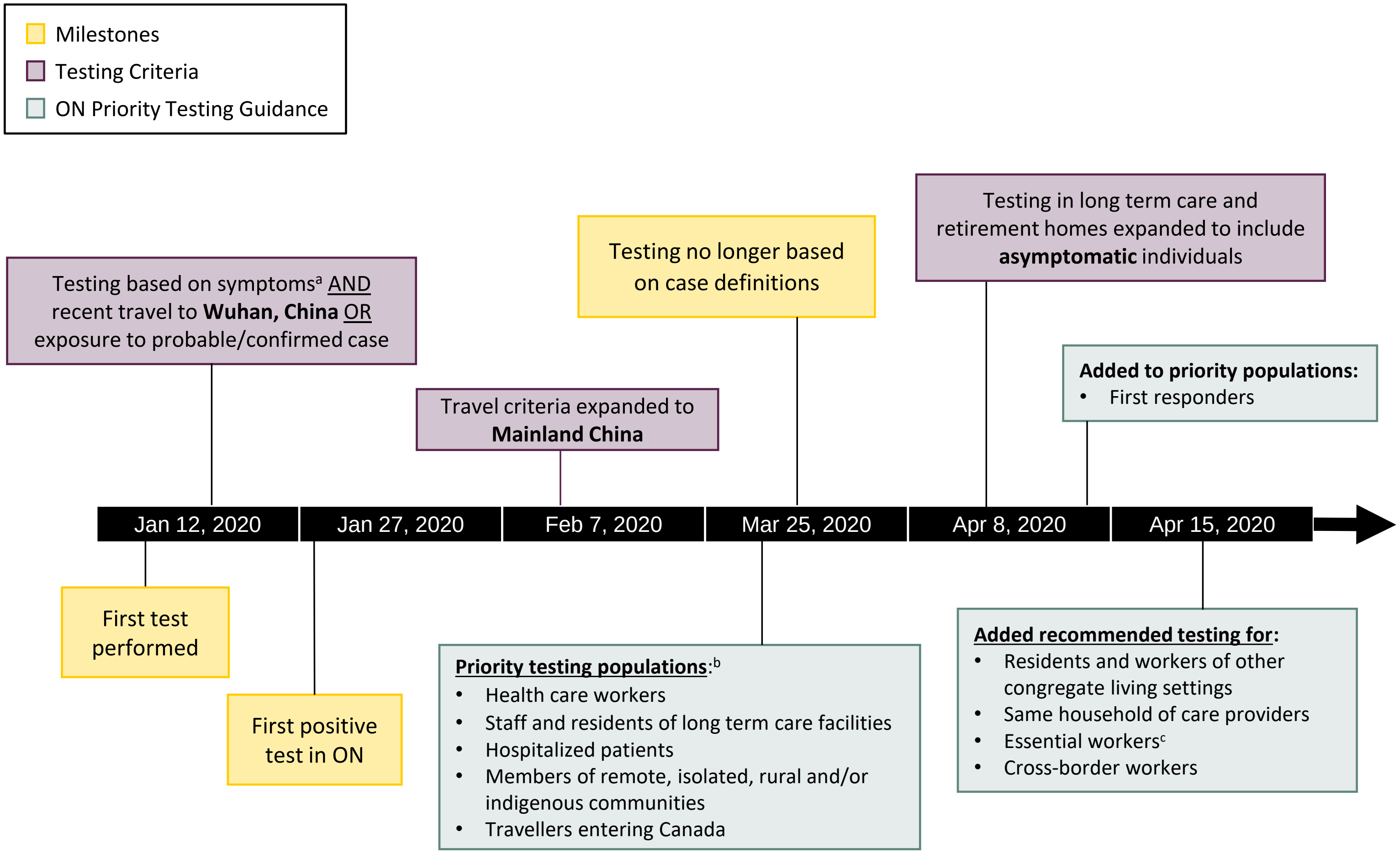

a. Fever and/or cough or difficulty breathing
b. Testing recommended if symptomatic (one of: fever, new/worsening symptom, clinical or radiologic evidence of pneumonia)
c. Provincial workers considered critical to preserving life, health and basic societal functioning - https://www.ontario.ca/page/list-essential-workplaces

**Figure S1.** Key dates related to SARS-CoV-2 testing in Ontario (ON) over the study period.

**Table S1.** Characteristics of the 118 individuals with invalid test results that did not have other specimens submitted to Public Health Ontario Laboratory, January 11, 2020 – April 22, 2020 with valid test results (positive, *n*=4; negative, *n*=23).

| Characteristics | Test[a] Result Invalid (*n*=118) |
|---|---|
| Age, y—median (IQR) | 44 (31–61) |
| Age group, y—No. (%) | |
| <1 | 0 (0.0) |
| 1–4 | 2 (1.7) |
| 5–19 | 8 (6.8) |
| 20–39 | 34 (28.8) |
| 40–59 | 38 (32.2) |
| 60–69 | 14 (11.9) |
| 70–79 | 10 (8.5) |
| 80–89 | 5 (4.2) |
| ≥90 | 6 (5.1) |
| Not specified | 1 (0.8) |
| Gender—No. (%)[b] | |
| Male | 53 (44.9) |
| Female | 64 (54.2) |
| Other/Not specified | 1 (0.8) |
| Reason for Testing[c] | |
| Symptoms[d]—No. (%) | |
| Yes | 106 (89.8) |
| No | 2 (1.7) |
| Not specified | 10 (8.5) |
| Travel history[e]—No. (%) | |
| Yes | 16 (13.6) |
| None reported | 102 (86.4) |
| Exposure history[f]—No. (%) | |
| Yes | 3 (2.5) |
| None reported | 115 (97.5) |
| Healthcare worker—No. (%) | 3 (2.5) |
| Ever hospitalized—No. (%) | 10 (8.5) |
| Outbreak associated—No. (%) | 4 (3.4) |
| Deceased (autopsy specimen)—No. (%) | 0 (0.0) |

*[a]Molecular-based testing included one or more of the following: end-point PCR + Sanger sequencing of the RNA-dependent RNA polymerase (RdRp) gene, rRT-PCR to detect the envelope (E) gene, RdRp gene and/or ORF1ab.*

*[b]Percentages have been rounded and may not total 100%.*

*[c]Reasons for testing were not mutually exclusive.*

*[d]Symptoms— Self-reported response or clinical assessment of fever, cough, sore throat, pneumonia, or other COVID-19 symptoms*

*[e]Travel history—Self-reported response to 'Travelled outside the country in the last 14 days?'.*

*[f]Exposure history—Self-reported response to 'Exposure to probable, or confirmed case?'.*

### APPENDIX I. Molecular-based Testing Supplementary

#### A.I.I. Supplementary Methods

##### *RNA extraction and molecular-based assays*

Automated processes were used for RNA extraction and included the NucliSENS easyMAG extractor (bioMérieux, QC, Canada), the *m*2000 platform (Abbott Inc., IL, USA), and the cobas® 8800 (Roche Diagnostics, Germany). All were used according to manufacturer instructions. For the Roche assay, a 400 µl aliquot of specimen was added to 200 µl of cobas® Omni Lysis Reagent (43% guanidine thiocyanate w/v). The instrument was loaded with a 400 µl aliquot of the above 600 µl combination of specimen/lysis reagent for testing.

The initial test for SARS-CoV-2 used easyMAG extracted RNA which was subjected to a nested end-point PCR[1] targeting a 192 base pair fragment of the SARS-CoV-2 RdRp gene, followed by Sanger sequencing using standard methods. Briefly, 5 µL extracted RNA was added to 20 µL of a reaction mixture containing 12.5 µL 2x Superscript-III One-step RT-PCR Master Mix, 1 µL BSA solution, $MgSO_4$ solution (5 nM), 1 µL Superscript-III RT enzyme solution, 3.1 µL of RNAse free and 1 µL each of 10 µM primers RdRp-PHL-WH-F (5'-TGCCATTAGTGCAAAGAATAGAGC-3') and RdRp-PHL-WH-R (5'-GCATGGCTCTATCACATTTAGG-3'). One-step RT-PCR amplification consisted of a pre-incubation at 50°C for 20 min and 94°C for 3 min, followed by initial denaturation at 95°C for 3 min, 45 cycles of denaturation at 95°C for 15 sec, annealing at 56°C for 15 sec, extension at 72°C for 30 sec and final extension at 72°C for 2 min. Next, 5 µL of the outer PCR product was added to 20 µL of a reaction mixture containing 12.5 µL Qiagen Taq PCR Master Mix, 3.1 µL of RNAse free and 0.4 µL each of 10 µM primers RdRp-PHL- WH-COV-3Fseq (5'-GCACCGTAGCTGGTGTCTCT-3'), and RdRp-PHL-WH-COV-3Rseq (5'-AATCCCAACCCATAAGGTGA-3'). Amplification of a 192 bp fragment was performed using the following cycling conditions: 95°C 3 min, 45 cycles at 95°C 30 sec, 56°C 30 sec, 72°C 30 sec, and a final extension at 72°C 10 min. The PCR product was purified sequenced using BigDye Terminator v3.1 Cycle Sequencing Kit (Applied Biosystems, Foster City) according to the manufacturer's instructions. Trimmed and cleaned sequencing results were aligned to the reference (GenBank accession MN908947) and a similarity of ≥99% was considered positive for SARS-CoV-2. Early specimens were also sent to Canada's National Microbiology Laboratory (NML) for secondary confirmation and supplementary testing using various NML-developed molecular-based assays.

Next, a laboratory developed test (LDT) rRT-PCR was used for detection of the envelope (E) gene and/or SARS-CoV-2 RNA-dependent RNA polymerase (RdRp) gene using simplex or duplex methods. Duplex rRT-PCR reactions combined the primers and probe for E-gene or RdRp gene with the internal control human RNase P (RNP), **Table S2**. Each 10 µl reaction contained 5 µl of eluted RNA, 2.5 µl of TaqPath™ 1-Step Multiplex Master Mix (Applied Biosystems), 0.4 µM each of forward and reverse primers and 0.2 µM of the labeled probe. Amplification and detection were performed using an ABI QuantStudio 5 Real-Time PCR System under the following conditions: 2 min at 25°C (uracil-DNA N-glycosylase incubation), 10 min at 53°C (reverse transcription), 95°C (polymerase activation) for 2 min, and 45 cycles of denaturation at 95°C for 3 sec and annealing/extension at 60°C (58°C for RdRp simplex) for 30 sec. Results of the LDT rRT-PCR were based on cycle threshold ($C_t$) values as follows: a $C_t$ ≤38 was categorized as detected (positive), 38.1 to 39.9 was indeterminate, and a $C_t$ ≥40 was not detected (negative). Valid results required a positive result for the RNP gene internal control. The testing algorithm initially required detection of both targets for laboratory confirmation. At first E-gene and RdRp were tested in simplex reactions on the same plate. After a period, with an increase in testing volumes and percent positivity, specimens were tested first for the E-gene target due to its greater sensitivity and at the end of the day in batches by RdRp for confirmation of E-gene-positive results. This was later revised to single target detection for COVID-19 diagnosis using the E-gene target.

As of April 3, 2020 an additional rRT-PCR assay using the cobas® 8800 (Roche) was brought online and used for detection of the SARS-CoV-2. The cobas® 8800 is an all-in-one system, which includes rRT-PCR detection of the E-gene and ORF1ab. Testing was performed according to manufacturer's instructions. Specimens tested in the cobas® SARS-CoV-2 rRT-PCR assay were reported as detected or not detected; the manufacturer does not include an indeterminate range. Although the $C_t$ value is obtainable from the instrument, the maximum number of cycles of PCR amplification used in the assay is proprietary and not provided in the kit insert and documentation.

An indeterminate result on a real-time PCR assay was defined as a late amplification signal in an rRT-PCR reaction at a predetermined high $C_t$ value range, which is delineated by the testing laboratory at the time of validation of a LDT, or by the manufacturer for commercial assays. This may be due to low viral target quantity in the clinical specimen approaching the limit of detection of the assay or alternatively, in rare cases, may represent nonspecific reactivity (false signal) in the specimen. When clinically relevant, repeat testing is recommended.

An uninterpretable result was classified as invalid, and reflected a failure in one or more of the test's internal controls. This may be due to any number of issues, such as the presence of PCR inhibitors or poor extraction due to a highly viscous specimen. Where it was not possible to repeat the test and obtain a valid result, the invalid result was reported as such.

**Table S2.** Primer and probe sequences[2] for detection of SARS-CoV-2 and the human RNAse P gene as an internal control[3], using rRT-PCR.

| Primers and Probes | Sequence (5' → 3') |
|---|---|
| SARS-CoV-2 Detection | |
| Envelope (E) gene | |
| E_Sarbeco_F1 | ACAGGTACGTTAATAGTTAATAGCGT |
| E_Sarbeco_R2 | ATATTGCAGCAGTACGCACACA |
| E_Sarbeco_P1 | FAM-ACACTAGCCATCCTTACTGCGCTTCG-BQH1 |
| RNA-dependent RNA polymerase (RdRp) | |
| RdRp_SARSr-F2 | GTGARATGGTCATGTGTGGCGG |
| RdRp_SARSr-R1 | CARATGTTAAASACACTATTAGCATA |
| RdRp_SARSr-P2 | FAM-CAGGTGGAACCTCATCAGGAGATGC-QSY |
| Human RNAse P (Internal Control) | |
| RNAse P gene | |
| RNase P-F | AGATTTGGACCTGCGAGCG |
| RNase P-R | GAGCGGCTGTCTCCACAAGT |
| RNP_JUN-QSY | JUN-TTCTGACCTGAAGGCTCTGCGCG-QSY |

##### *Statistical Analysis*

We measured correlations between $C_t$ values of the E-gene and RdRp targets (LDT), and the E-gene and ORF1ab (Roche) using Pearson correlation coefficients. For $C_t$ comparisons of dual targets within the same assay, the $C_t$ for negative results was set to 40 to not bias single target positive results.

#### A.I.II. Supplementary Test Performance Characteristics

The majority (99.0%, *n*=86,071) of specimens were tested with a single assay, the largest number of which were tested using PHO's LDT rRT-PCR assay (*n*=56,928) or the Roche platform (*n*=29,141), **Table S3**.

**Table S3.** Number of specimens tested by a single assay and with an additional assay(s): end-point RdRp gene PCR with Sanger sequencing (End Point), referred testing to the National Microbiology Laboratory (Referred), rRT-PCR laboratory developed test (LDT), rRT-PCR Roche cobas® 8800 (Roche), *n*=86,942.

| | Initial Test | | | |
|---|---|---|---|---|
| Additional Test(s) | End-Point | Referred | Lab Developed | Roche |
| None | 1 | 1 | 56,928 | 29,141 |
| +Lab Developed | 45 | 410 | – | 23 |
| +Referred | 370 | – | 0 | 0 |
| +Lab Developed +Reference | 23 | 0 | 0 | 0 |

Of the 871 specimens that were tested using multiple platforms, 24 (2.8%) specimens had an invalid or indeterminate result for one of the tests. Concordant results were observed for 831 (95.4%) specimens tested using multiple assays. Of the 23 specimens with discordant results, 19 were related to negative end-point PCR results, and where rRT-PCR had also been performed a median of $C_t$ 36.2 (IQR 34.4–36.9) was observed. Details of the discordant results can be found in **Tables S4**.

**Table S4.** Number of specimens by discordant result where tested by end-point RdRp PCR with Sanger sequencing (End-Point), referred testing to the National Microbiology Laboratory (Referred) (*n*=23), and of these 17 were additionally tested by rRT-PCR laboratory developed test (LDT).

| End-Point | Referred | Lab Developed, Median $C_t$ (IQR) | No. Specimens |
|---|---|---|---|
| Negative | Negative | Indeterminate, 39.5 | 1 |
| Negative | Negative | Positive, 35.7 (34.3–37.2) | 6 |
| Negative | Indeterminate | Positive, 36.2 (35.4–36.8) | 5 |
| Negative | Positive | Positive 32.2 (29.8–34.5) | 2 |
| Negative | Positive | – | 5 |
| Indeterminate | Negative | Positive, 36.1 | 1 |
| Positive | Negative | Positive, 34.4 | 1 |
| Positive | Negative | – | 1 |
| Positive | Positive | Negative | 1 |

*Abbreviations: Ct, cycle threshold; IQR, interquartile range.*

##### *rRT-PCR assays and targets*

To explore the performance of the different rRT-PCR targets, we compared test results using one or both targets across different assays. Of the 57,429 specimens tested by LDT, 3,717 (6.5%) were positive for either E-gene or RdRp-gene (**Table S5**) – the majority of which was E-gene alone (78.1%). Where both targets were tested for a specimen (*n*=12,592), 1,077 (8.6%) were positive for both E- and RdRp-genes, 94 (0.7%) were E-gene positive only, and two specimens were indeterminate for the E-gene with one negative and one positive for RdRp. Of the 29,164 specimens tested using the Roche platform, 3,788 (13.0%) were positive for E-gene and/or ORF1ab gene, 3,507 (12.0%) were positive for both targets, 225 (0.8%) were positive for E-gene only, and 56 (0.2%) were positive for ORF1ab only. There were 7 (0.02%) specimens with invalid results which subsequently tested negative using the LDT assay.

**Table S5.** Number of specimens by rRT-PCR assay result for laboratory developed test (LDT) envelope (E-gene) and RdRp targets and Roche envelope (E-gene) and ORF1ab target.

| Assay Results | E-gene Results | | |
|---|---|---|---|
| Second Target | Positive | Negative | Indeterminate |
| LDT[a] | | | |
| RdRp results | | | |
| Positive | 1,077 | 0 | 1 |
| Negative | 94 | 11,419 | 1 |
| Indeterminate | 0 | 0 | 0 |
| Not done | 2,546 | 42,270 | 19 |
| Roche[b] | | | |
| ORF1ab results | | | |
| Positive | 3,507 | 56 | – |
| Negative | 225 | 25,369 | – |

*Abbreviations: IQR, interquartile range; Max, maximum; Min, minimum.*
*[a]Invalid results, n=2.*
*[b]Invalid results, n=7.*

We compared $C_t$ values (where available) for each specimen tested using multiple targets on the same platform, which revealed highly correlated $C_t$ values between targets (LDT $R^2$=0.91; Roche $R^2$=0.98), **Figure S2**. However, we noted that when comparing results of the two targets tested for the same specimen the LDT E-gene $C_t$ was lower than the RdRp-gene $C_t$ for 896 (77.4%) of 1,158 specimens (**Table S6**), with a median E-gene $C_t$ of 22.4 (IQR 18.3–27.0), and a median RdRp $C_t$ of 23.2 (IQR 19.5–27.5), $p$<0.001. In contrast, the Roche E-gene $C_t$ was higher than the ORF1ab $C_t$ for 3,339 (88.6%) of 3,768 specimens (**Table S6**), with a median E-gene $C_t$ of 26.7 (IQR 21.0–33.7) and a median ORF1ab $C_t$ of 26.2 (IQR 20.6–31.9), $p$<0.001. Although the largest differences in $C_t$ values for the two the targets on the same platform was up to 16.4 and 17.1 for the LDT and Roche platforms, respectively, it should be noted that the median differences were less than two $C_t$. While the median $C_t$ for the E-gene on the Roche platform was slightly higher compared to ORF1ab, it is a more sensitive target as more specimens were E-positive/ORF1ab-negative (*n*=225) than ORF1ab-positive/E-negative (*n*=56), **Table S5**.

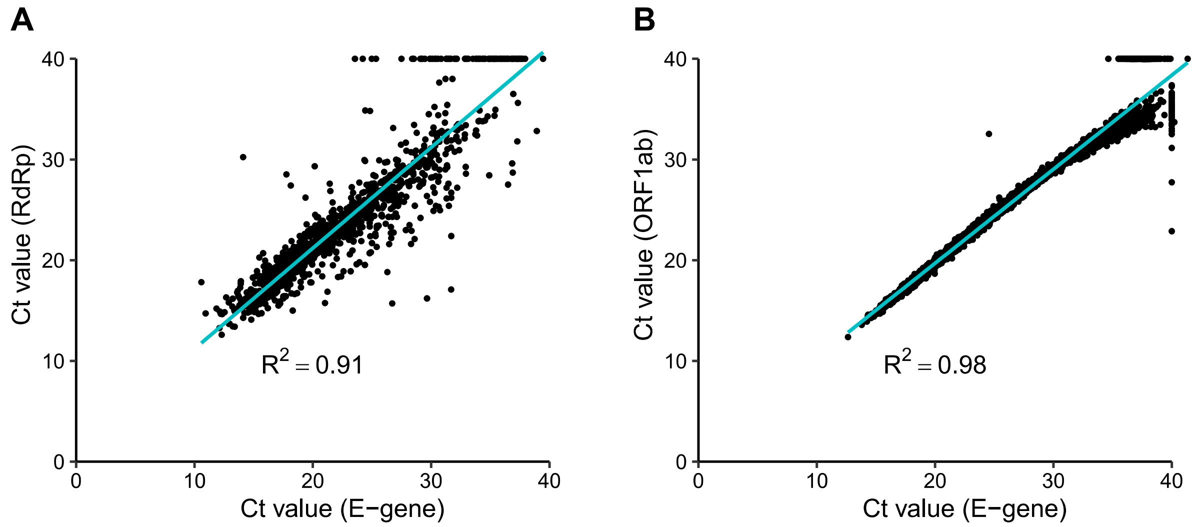

**Figure S2.** Scatter plot of cycle threshold ($C_t$) values for tests with multiple targets. **(A)** Laboratory developed test targets, envelope gene (E-gene) and RdRp, and **(B)** Roche targets, envelope gene (E-gene) and ORF1ab. $C_t$ values were set to 40 for a negative test result where the other target was positive to not bias the results towards those where both targets were positive.

**Table S6.** Relative difference in cycle threshold ($C_t$) for specimens tested using rRT-PCR for both the envelope (E-gene) and RdRp targets by LDT (*n*=1,158), or tested for both the envelope (E-gene) and ORF1ab targets by the Roche assay (*n*=3,768). E-gene $C_t$ was categorized as higher or lower than the second target $C_t$. $C_t$ values were set to 40 for a negative test result where the other target was positive to not bias the results towards those where both targets were positive.

| | | $C_t$ value Relative Difference | |
|---|---|---|---|
| Category | *n* (%) | Median (IQR) | Min–Max |
| LDT | | | |
| E-gene $C_t$ lower than RdRp $C_t$ | 896 (77.4) | 1.5 (0.8–2.5) | 0.01–16.44 |
| E-gene $C_t$ higher than RdRp $C_t$ | 253 (21.8) | 1.1 (0.4–2.4) | 0.03–14.60 |
| Identical $C_t$ | 9 (0.8) | 0.0 (0.0–0.0) | 0.00–0.00 |
| Roche | | | |
| E-gene $C_t$ lower than ORF1ab $C_t$ | 413 (11.0) | 1.5 (0.1–2.5) | 0.01–8.00 |
| E-gene $C_t$ higher than ORF1ab $C_t$ | 3,339 (88.6) | 0.7 (0.3–1.5) | 0.01–17.12 |
| Identical $C_t$ | 16 (0.4) | 0.0 (0.0–0.0) | 0.00–0.00 |

*Abbreviations: IQR, interquartile range; Max, maximum; Min, minimum.*

##### A.I.III. Supplementary Molecular-based Testing Summary

Examining the different targets and assays used over the first few months of the COVID-19 pandemic showed good correlation between targets within an assay; however, we found that percent positivity and median $C_t$ varied across different assays. This can largely be explained by a bias in assays used based on the swab manufacturer (e.g. Roche swabs used with the cobas® 8800 platform are distributed by PHO largely to hospitals), and the timeline of these assays coming online in the context of the prioritized population for testing and case rate at the time.

#### APPENDIX II. Specimen-based Testing Supplementary

**Table S7.** Summary for the number of specimens per individual received by Public Health Ontario, January 11, 2020 – April 22, 2020.

| Number of Submitted Specimens | Number of Individuals (%) |
|---|---|
| 1 | 74,779 (93.1) |
| 2 | 4,837 (6.0) |
| 3 | 568 (0.7) |
| 4 | 126 (0.2) |
| 5+ | 44 (0.1) |

**Table S8.** Number of same-day multi-specimen testing episodes categorized by SARS-CoV-2 molecular-based[a] test result and specimen types.

| | Number of Testing Episodes by Results | | |
|---|---|---|---|
| Specimen Types[b] | All Positive (*n*=105) | All Negative (*n*=2,882) | Positive and Negative or Other[c] Discordant (*n*=37) |
| Exclusively Upper Respiratory | | | |
| NPS only | 23 | 311 | 12 |
| NPS + TS | 68 | 2,396 | 23[d] |
| NPS + Other | | 2 | |
| TS only | 1 | 8 | |
| TS + Other | | 17 | |
| Other only | | 2 | |
| Exclusively Lower Respiratory | | | |
| BAL only | | 10 | |
| BAL + Other | | 2 | |
| Sputum only | | 1 | |
| Other only | | 6 | |
| Upper and Lower Respiratory | | | |
| NPS + BAL | | 3 | |
| NPS + Sputum | 3 | 28 | 1 |
| NPS + TS + Sputum | | 28 | |
| NPS + TS + BAL | | 1 | |
| NPS + TS + Other LResp | 1 | 1 | |
| NPS + TS + Sputum + Other UResp | | 1 | |
| NPS + Other LResp + Other UResp | | 1 | |
| NPS + Other LResp | | 9 | |
| TS + Sputum | 1 | 2 | |
| Sputum + Other UResp | | 1 | |
| Non-Respiratory | | | |
| Non-Respiratory only | | 1 | |
| NPS + TS + Non-Respiratory | | 1 | |
| Not Specified | | | |
| NPS + TS + Not Specified | 2 | 3 | |
| NPS + TS + Other UResp + Not Specified | | | 1 |
| NPS + TS + Other LResp + Not Specified | 1 | | |
| NPS + TS + BAL + Not Specified | | 1 | |
| NPS + Not Specified | 2 | 13 | |
| TS + Not Specified | 1 | 8 | |
| Sputum + Not Specified | | 1 | |
| Not Specified only | 2 | 24 | |

*Abbreviations: BAL, bronchoalveolar lavage; LResp, lower respiratory; NPS, nasopharyngeal swab, TS, throat swab, UResp, upper respiratory.*

[a]*Molecular-based testing included one or more of the following: end-point PCR + Sanger sequencing of the RNA-dependent RNA polymerase (RdRp) gene, rRT-PCR to detect the envelope (E) gene, RdRp gene and/or ORF1ab.*

[b]*Specimen category "other" consists of specimens such as oral swab, saliva, tracheal, lung tissue.*

[c]*Category includes NPS-positive/TS-indeterminate result (n=1), and NPS-negative/NPS-indeterminate result (n=1).*

[d]*Comprised of 17 NPS-positive/TS-negative, 6 NPS-negative/TS-positive and 1 NPS-positive/TS-indeterminate.*

**Table S9.** Number and proportion of individuals by symptom status at time of testing, for those initially testing negative and on subsequent test was either positive ≤7 days following initial negative result, or positive >7 days following initial negative.

| Initial Negative Test[a] | Subsequent Positive Test[a] *n* (%)[b] Symptoms (subsequent positive ≤ 7 days) | | | | Symptoms (subsequent positive > 7 days) | | | |
|---|---|---|---|---|---|---|---|---|
| | Yes | No | Not Stated | Total | Yes | No | Not Stated | Total |
| Symptoms | | | | | | | | |
| Yes | 39 (69.6) | 0 (0.0) | 17 (30.4) | 56 (50.9) | 63 (59.4) | 3 (2.8) | 40 (37.7) | 106 (73.1) |
| No | 5 (83.3) | 0 (0.0) | 1 (16.7) | 6 (5.5) | 5 (83.3) | 1 (16.7) | 0 (0.0) | 6 (4.1) |
| Not Stated | 30 (62.5) | 1 (2.1) | 17 (35.4) | 48 (43.6) | 22 (66.7) | 1 (3.0) | 10 (30.3) | 33 (22.8) |

[a]*Molecular-based testing included one or more of the following: end-point PCR + Sanger sequencing of the RNA-dependent RNA polymerase (RdRp) gene, rRT-PCR to detect the envelope (E) gene, RdRp gene and/or ORF1ab.*
[b]*Percentages have been rounded and may not total 100%.*

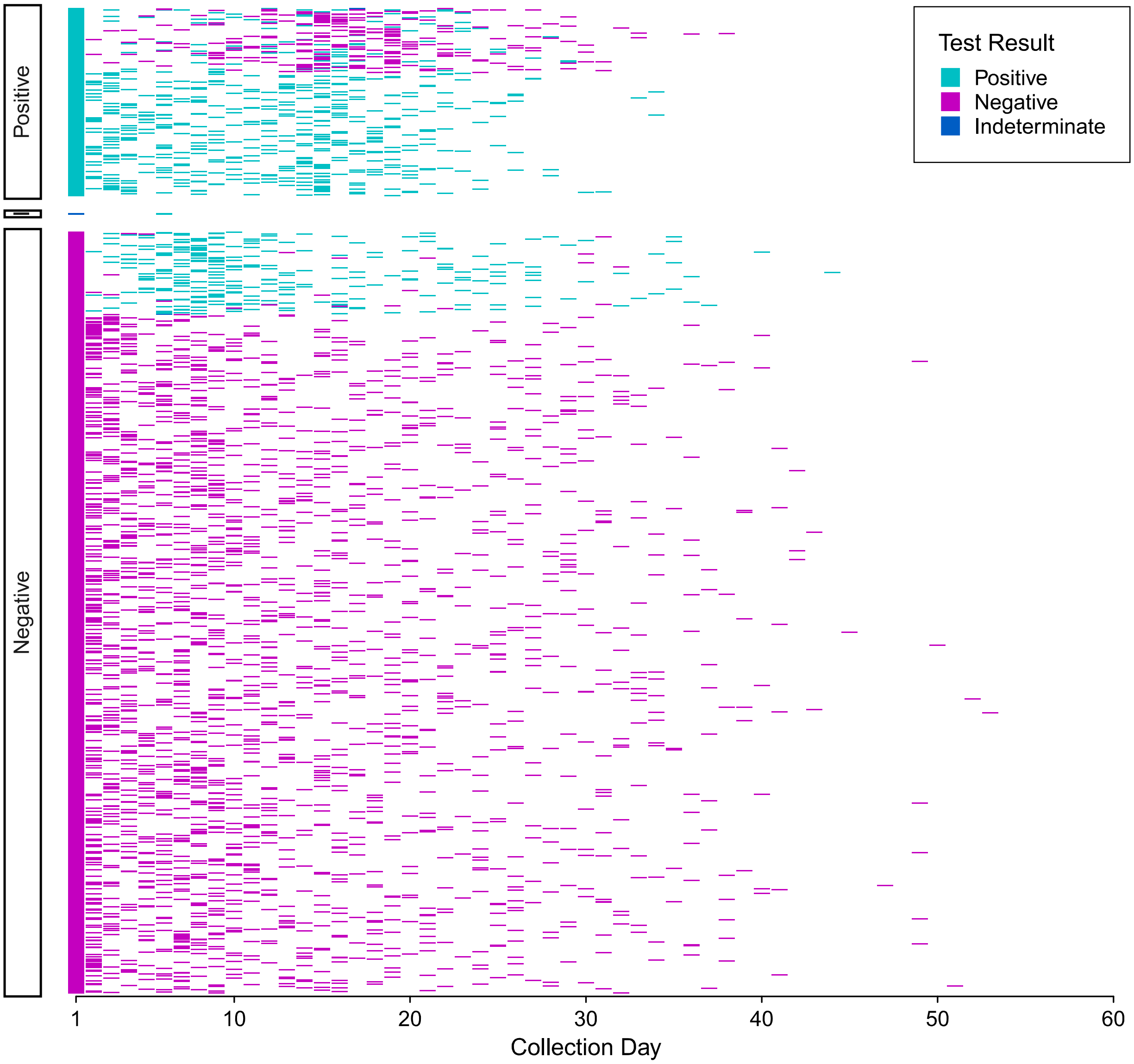

**Figure S3.** Molecular-based test results for individuals with specimens collected on more than one day and submitted for testing to Public Health Ontario (*n*=2,943). Divided by initial (day 1) test result, positive, indeterminate (I) or negative, serial test results for each individual are represented horizontally across collection days. Any positive result for an individual with multiple specimens collected on the same-day is displayed as positive, and indeterminate where there was an indeterminate result and no positive. Testing included one or more of the following: end-point PCR + Sanger sequencing of the RNA-dependent RNA polymerase (RdRp) gene, rRT-PCR to detect the envelope (E) gene, RdRp gene and/or ORF1ab.
